# The impact of social distancing and epicenter lockdown on the COVID-19 epidemic in mainland China: A data-driven SEIQR model study

**DOI:** 10.1101/2020.03.04.20031187

**Authors:** Yuzhen Zhang, Bin Jiang, Jiamin Yuan, Yanyun Tao

## Abstract

The outbreak of coronavirus disease 2019 (COVID-19) which originated in Wuhan, China, constitutes a public health emergency of international concern with a very high risk of spread and impact at the global level. We developed data-driven susceptible-exposed-infectious-quarantine-recovered (SEIQR) models to simulate the epidemic with the interventions of social distancing and epicenter lockdown. Population migration data combined with officially reported data were used to estimate model parameters, and then calculated the daily exported infected individuals by estimating the daily infected ratio and daily susceptible population size. As of Jan 01, 2020, the estimated initial number of latently infected individuals was 380.1 (95%-CI: 379.8∼381.0). With 30 days of substantial social distancing, the reproductive number in Wuhan and Hubei was reduced from 2.2 (95%-CI: 1.4∼3.9) to 1.58 (95%-CI: 1.34∼2.07), and in other provinces from 2.56 (95%-CI: 2.43∼2.63) to 1.65 (95%-CI: 1.56∼1.76). We found that earlier intervention of social distancing could significantly limit the epidemic in mainland China. The number of infections could be reduced up to 98.9%, and the number of deaths could be reduced by up to 99.3% as of Feb 23, 2020. However, earlier epicenter lockdown would partially neutralize this favorable effect. Because it would cause in situ deteriorating, which overwhelms the improvement out of the epicenter. To minimize the epidemic size and death, stepwise implementation of social distancing in the epicenter city first, then in the province, and later the whole nation without the epicenter lockdown would be practical and cost-effective.

## Introduction

The coronavirus disease 2019 (COVID-19), initially taken as “pneumonia of unknown etiology”, emerged in December 2019, Wuhan, Hubei Province, China. The causative pathogen was announced by the Chinese Center for Disease Control and Prevention (China CDC) on Jan 08, 2020, to be a novel coronavirus [1], lately named severe acute respiratory syndrome coronavirus 2 (SARS-CoV-2)[2]. COVID-19 broke out in Wuhan in January 2020, and spread to the whole Hubei Province, the rest of China and abroad with astonishing speed. On Jan. 31, 2020, the World Health Organization (WHO) announced that COVID-19 constitutes a “public health emergency of international concern”. As of Feb 28, there were 7,8961 cases confirmed in China, and 4,691 cases confirmed in 51 other countries. On that day, WHO increased the assessment of the risk of spread and risk of impact of COVID-19 to very high at the global level[3].

Apart from the intrinsic infectivity of the virus, population mobility and epidemic prevention and control measures could affect the prevalence scale. Unfortunately, the prevalence of COVID-19 encountered the Spring Festival Migration of China, the world’s largest annual human migration as hundreds of millions of people rush home for family reunions. In addition, the epicenter Wuhan is the capital of Hubei Province of China. It has a population of more than 15 million, including resident and floating population, and it situates at the transportation hub in the central China area. As of Jan 23, 2020, more than 5 million people were migrating out of Wuhan according to official accounts [3]. Emergency monitoring and close contact management in Wuhan was carried out since Jan 03, 2020; China CDC Level 2 emergency response activated on Jan 06, and Level 1 emergency response activated on Jan 15 [1]. On Jan 20, 2020, COVID-19 was included in the statutory report of Class B infectious diseases, managed as Class A infectious diseases by the National Health Commission of China. The Chinese government locked down the Wuhan city on Jan 23, and then locked down other cities of Hubei Province immediately after. By Jan 25, 30 provincial governments in China activated first-level public health emergency response. Hence, In addition to strict quarantine management, substantial social distancing measures to limit population mobility and to reduce within-population contact rates were executed almost in the whole country. For example, public activities were canceled; communities adopted enclosed management; the national holiday of Spring Festival and the winter vacation were extended so that work resumption and school re-opening could be extensively postponed. In addition, people were required to wear facemasks in public. However, with all those efforts, the prevalence of COVID-19 was escalating.

Epidemic prevention and control strategies need to be re-examined. Vaccine and antiviral drug development is the ultimate way to defeat a virus, but it is time-consuming. Non-pharmaceutical interventions to interrupt transmission could be implemented immediately, gaining time for pharmaceutical development. Briefly, there were three steps of non-pharmaceutical interventions for reducing contact rates between susceptible individuals and infected individuals. First, quarantine management, i.e. quarantining the infected, the suspicious and their close contacts; second, social distancing to confine within-population contact; third, locking down the epicenter to prevent further exportation of infected and latently infected individuals to other regions.

Quarantine management is a fundamental measure ought to be taken once the human-human transmission is confirmed. Theoretically, if substantial social distancing and/or epicenter lockdown were implemented early enough, there would be no prevalence or no spreading. But realistically, it takes time for preliminary investigation. Besides, rigorous measures would bring about deep social influences and economic consequences. So, it is challenging to choose the right response at the right scale in the right area at the right time[4], especially when the transmission pattern and clinical characteristics were not fully understood.

The importance of non-pharmaceutical control measures requires further research to quantify their impact [3]. Mathematical models are useful to evaluate the possible effects on epidemic dynamics of preventive measures, and to improve decision-making in global health [5,6]. In this study, we developed data-driven susceptible-exposed-infectious-quarantine-recovered (SEIQR) models to simulate the epidemic under the interventions of social distancing and epicenter lockdown, and to evaluate the impact of earlier interventions on the epidemic size and death number totally and respectively in Wuhan, Hubei Province, and 12 other provinces and municipalities. Population migration data combined with officially reported data were used to estimate model parameters, the number of the latently infected individuals, and daily exportation of the infected and latently infected from Wuhan. Our work provided evidence for the decision-making concerning prevention and control of the ongoing COVID-19 epidemic in other countries and future infectious disease epidemic.

### Data source and framework of the estimation model

In this study, SEIQR models were developed to simulate the COVID-19 epidemic in China. We built three SEIQR models to simulate the epidemic in Wuhan, the rest of Hubei Province, and other provinces and municipalities in China, respectively. Other provinces and municipalities studied here include nine provinces (i.e. Zhejiang, Guangdong, Henan, Hunan, Anhui, Jiangxi, Jiangsu, Shangdong and Sichuan) and three municipalities (i.e. Chongqing, Beijing and Shanghai) in China, which covered 90% infections in mainland China. For convenience, “Hubei” represents the rest of Hubei Province excluding Wuhan, and “other provinces” represents the above-mentioned provinces and municipalities in the text.

Figure 1 describes the framework of building estimation models. The reported confirmed, recovered and death cases were used to build initial models for Wuhan, Hubei and other provinces. Since there could be considerable under-reporting bias in the epicenter for lots of reasons. We assumed that the cases reported by other provinces were more accurate. Therefore, the model built on the cases reported by other provinces could estimate more accurately the number of infections. The reproductive number, infectious period and mean latent of models built for Wuhan and Hubei should be revised based on the latently infected individuals. The number of latently infected individuals in Wuhan before Jan 23 was estimated by the latently infected ratio and the daily population migration data from Wuhan to the rest of China. In the revised models of Wuhan and Hubei, parameters involving social distancing measures such as the removed rate, the decline factor of the reproductive number and the isolating ratio of susceptible individuals inherited from the initial models. The daily latent infected ratio is calculated based on the daily number of exposed individuals in Wuhan and the daily susceptible population size. Through the daily latent infected ratio and the daily number of individuals leaving Wuhan, the daily number of exposed individuals exported to Hubei and other provinces can be estimated.

**Figure 1.**
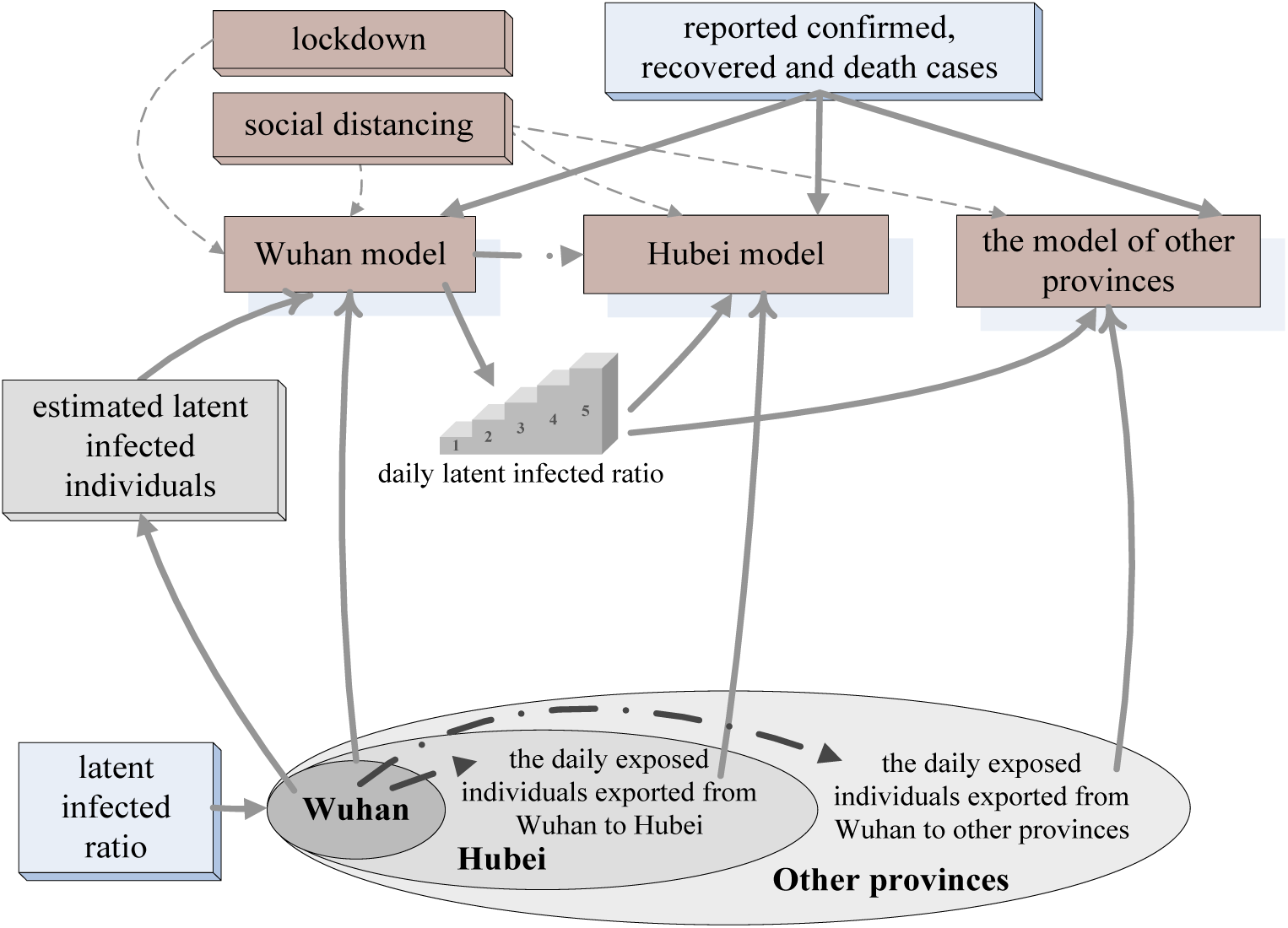
The framework of building models for Wuhan, Hubei and other provinces. We added the intervention measures, i.e. social distancing and epicenter lockdown, to models. These models were able to simulate the COVID-19 epidemic with interventions activated on different dates.

Wuhan has a population of around 15.39 million, including a resident population of 9 million, and nearly 6.39 million floating population exported before Jan 23 [7]. The population exported from Wuhan is the main source of imported cases in the rest of China. The travelers flowed from the rest of Hubei to other provinces, between two of the other provinces, and heading to Wuhan were not considered in this study. We assumed that the population exported from Wuhan was susceptible and exposed population. During the period of time from Jan 01 to Jan 22, 2020, the number of exported population from Wuhan was derived from two sources, a) domestic passenger number by air, train and road provided by Cao, Z., et al [7], and b) international outbound passenger number by air departed from Tianhe Airport (Wuhan, China) provided by a commercial APP named “Flight Steward”. Officially reported case data by the National Health Commission were used to estimate the parameters of SEIQR models.

As of Dec 31, 2019, there were 47 confirmed COVID-19 cases in Wuhan [1]. We took that 47 cases as the initial number of infected individuals in the model. From Jan 1 to 23, 2020, the estimated latent infected ratio in Wuhan is 0.12% [7]. As of Jan 23, the accumulated number of latently infected individuals in Wuhan and exported infected individuals estimated by the latently infected ratio were 10800 and 7677.6, respectively.

## Results

The estimated number of latently infected individuals by Jan 01, 2020 in Wuhan was 380.9 (95%-CI: 379.8∼381.0). The estimated base population size of daily susceptible individuals was 2*106. The base reproductive number r0 of Wuhan and Hubei was set to 2.2 (95%-CI: 1.4∼3.9) referring to the study of Liu, Q., et al [1]. When the interventions continue for 30 days, *r*_0_ was reduced to 1.58 (95%-CI: 1.34∼2.07). In other provinces, the estimated average base reproductive number *r*_0_ was 2.52 (95%-CI: 2.43∼2.63), and with 30-day interventions, *r*_0_ was reduced to 1.65 (95%-CI: 1.63∼1.69). The estimated infectious period *T*_I_ was 2.26 days (95%-CI: 2.14∼2.39) in Wuhan and Hubei, and was 3.75 days (95%-CI: 3.43∼4.13) in other provinces. The estimated mean latent was 10.1 days (95%-CI: 8.82∼11.78) in Wuhan and Hubei, and was 11.01 days (95%-CI: 9.51∼13.06) in other provinces. The estimated removed rate was 0.0125 (95%-CI: 0.008∼0.017) in Wuhan, 0.0175 (95%-CI: 0.0126∼0.0224) in Hubei, and 0.0185 (95%-CI: 0.014∼0.023) in other provinces. The initial ratio *λ*_0_ of isolating susceptible individuals was 0.1108 (95%-CI: 0.0957∼0.1254) in Wuhan, and 0.2326 (95%-CI: 0.2125∼0.2538) in Hubei, both of which were much lower than 0.4113 (95%-CI: 0.3794∼0.4432) in other provinces.

We investigated the estimated epidemic size when social distancing activated on different dates in different areas and at different strength levels in Figure 2. The results in Figure 2 (a) demonstrated that earlier intervention of social distancing in Wuhan could reduce the number of infections, but was not significant enough. Earlier intervention simultaneously in Wuhan and Hubei could further reduce the epidemic size. Activating at earlies time in Wuhan could allow Hubei activated later with an acceptable compromise. In Figure 2 (b), earlier intervention in other provinces could reinforce the effect of earlier intervention in Wuhan and Hubei. In order to control the epidemic size smaller than that of severe acute respiratory syndrome (SARS) in mainland China 2003 (5,779), social distancing should be activated simultaneously in Wuhan and Hubei on Jan 03, and in other provinces before Jan 15, or activated in Wuhan on Jan 03, in Hubei on Jan 6, and in other provinces before Jan 11. However, it could not be controlled smaller than the global size of the Middle East Respiratory Syndrome (MERS) epidemic 2015 (2,269). Of note, if the social distancing in Hubei lagged behind Wuhan for more than two weeks, the earlier intervention in other provinces could barely affect the epidemic size in China. Figure 2 (c) showed that, without social distancing (0\**λ*_0_), the infection number would be remarkably increased than the current epidemic estimation. Earlier activation of social distancing at low strength level (0.25\**λ*_0_) could be a bit better, but may eventually, maybe after Apr 1, enlarge the epidemic size. Earlier activation of social distancing at a moderate level (0.50\**λ*_0_ and 0.75\**λ*_0_) would be effective. Reasonably, earlier activating substantial social distancing at high-level (1\**λ*_0_), which is the actual social distancing strength implemented in China, could lead to the best results.

**Figure 2.**
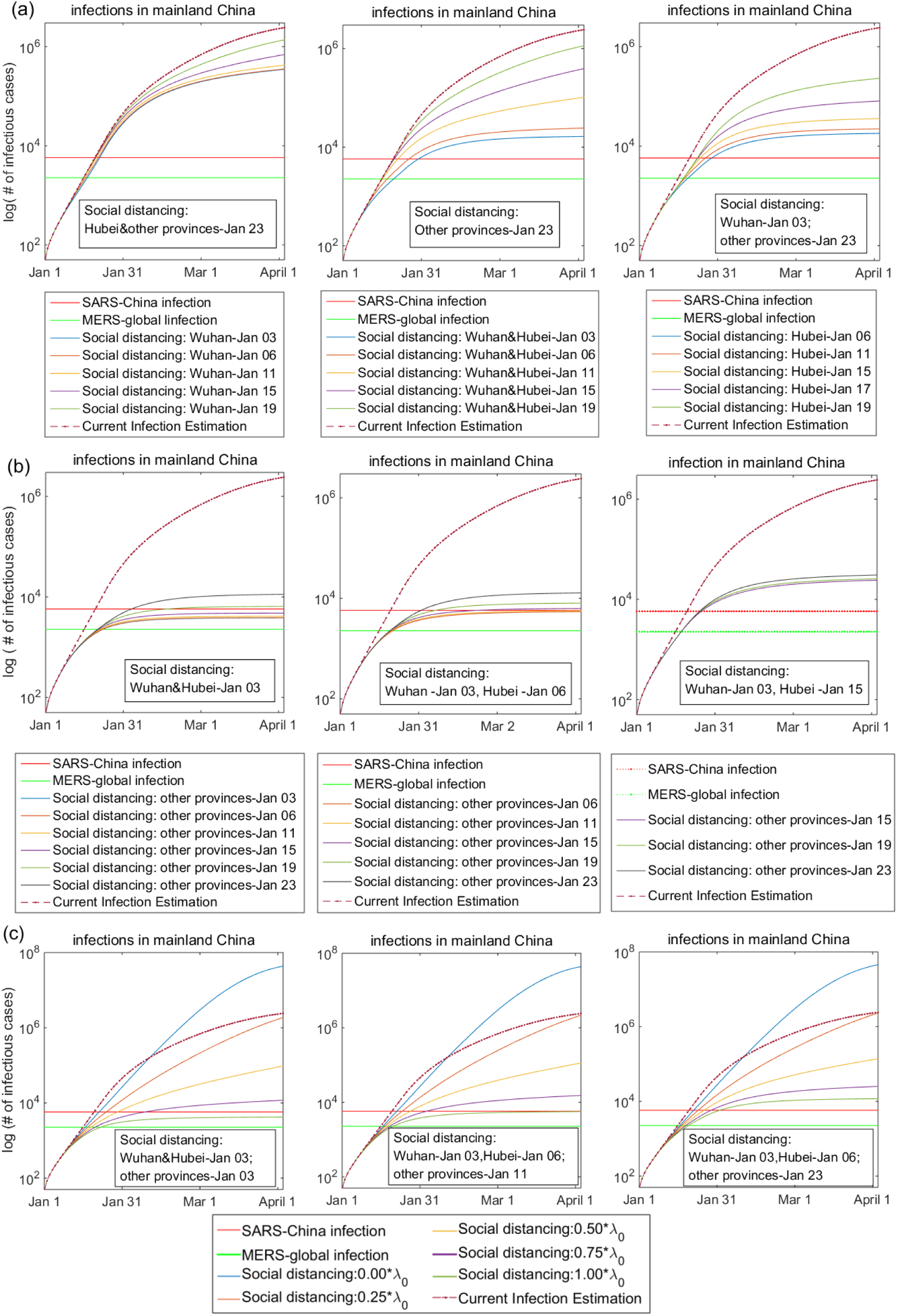
The estimated epidemic size when social distancing activated on different dates in different areas and at different strength levels. (a) The impact of different timing to activate social distancing in Wuhan and Hubei on the epidemic size, while activated on Jan 23 in other provinces; (b) The impact of different timing to activate social distancing in other provinces, while activated on Jan 3 in Wuhan unchangingly, and on Jan 3, 6 and 15 in Hubei respectively. (c) The impact of social distancing at different strength levels (i.e. *λ=* 0.00\**λ*_0_, 0.25\**λ*_0_, 0.50\**λ*_0_, 0.75\**λ*_0_ and 1.00\**λ*_0_) on the epidemic size. Current infection estimation: the estimated number of infection cases under current strategies, i.e. social distancing implemented in Wuhan, Hubei and other provinces simultaneously on Jan 23. SARS-China infection: the infection number of the severe acute respiratory syndrome (SARS) in mainland China 2003, that is 5,779. MERS-global infection: the global infection number of Middle East Respiratory Syndrome (MERS) 2015, that is 2,269. 0.25\**λ*_0_: 25% of the actual social distancing strength implemented in China. 0.50\**λ*_0_: 50% of actual strength. 0.75*λ_0:_ 75% of the actual strength. 1.00*λ_0_: the actual social distancing strength implemented in China.

We next investigated the impact of earlier epidemic lockdown on the estimated epidemic size and the number of deaths in Figure 3. The number of deaths was estimated by the number of infections multiplied with the current death rate in Wuhan, Hubei and other provinces (i.e. 4.43%, 3.09% and 0.835%). Unexpectedly, we found that the significant decrease of the nationwide epidemic size and death number by earlier social distancing would be partially neutralized by earlier epicenter lockdown (Figure 3a). With earlier social distancing in three different timings, the later of the lockdown, the smaller of the nationwide epidemic size and the less number of deaths there would be. This tendency is especially remarkable when social distancing was activated at the earliest time. With most strategies discussed here, the death number of COVID-19 could be controlled less than that of SARS in mainland China (829). Earlier social distancing without earlier lockdown could control the death number less than the global death number of MERS 2015 (431). Further investigation suggested that earlier epicenter lockdown would increase the number of infections and deaths in the epicenter Wuhan, while reducing the number of infections and deaths in the rest of Hubei and other provinces.

**Figure 3.**
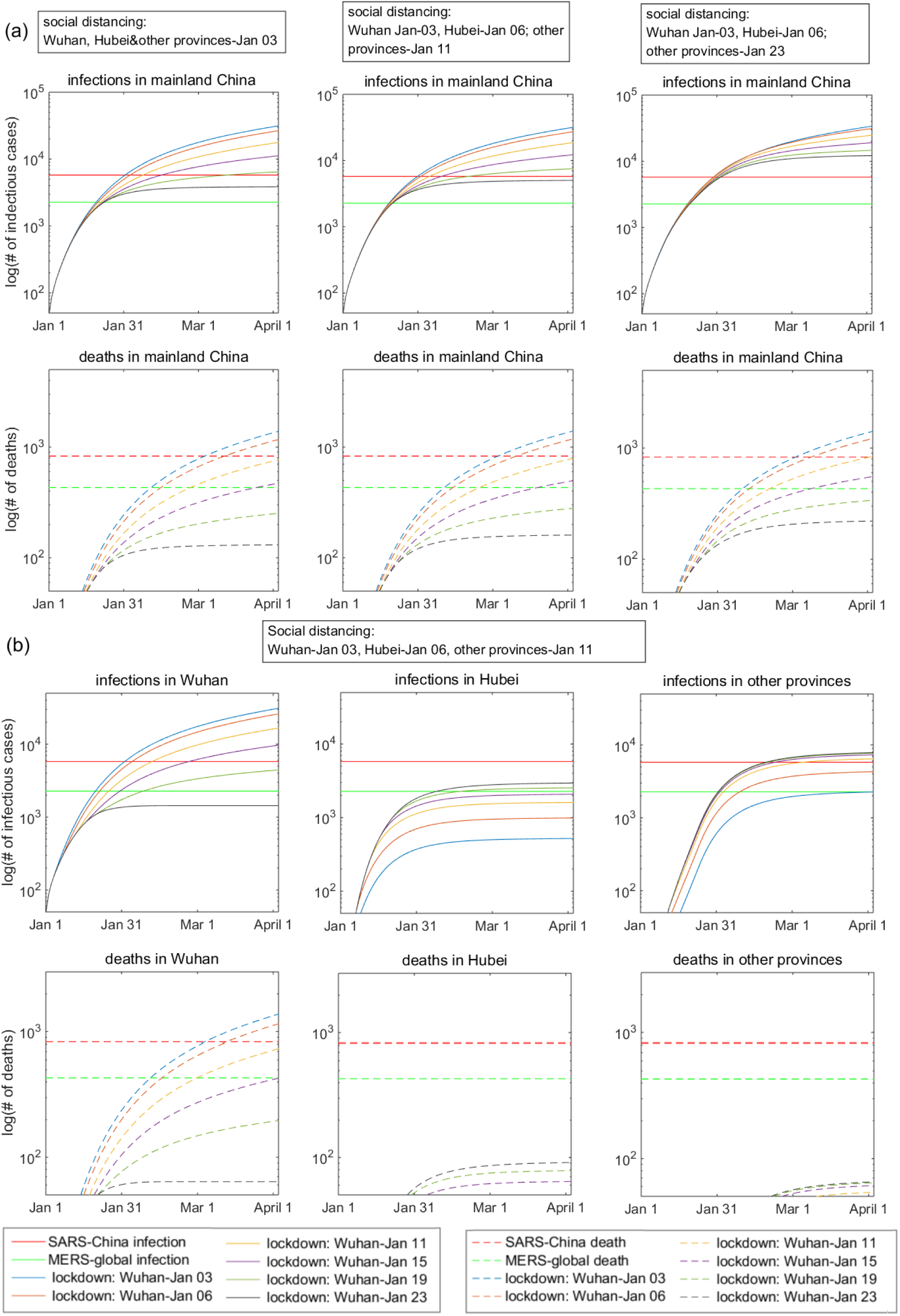
The estimated epidemic size and the number of deaths if Wuhan was put on lockdown before Jan 23. (a) The impact of the different timing of epidemic lockdown on the number of infections (upper row) and deaths (lower row) in mainland China with three different social distancing timings. Left column: social distancing activated in the whole nation simultaneously on Jan 03. Middle column: social distancing activated on Jan 03 in Wuhan, on Jan 06 in Hubei, and on Jan 11 in other provinces. Right column: social distancing activated on Jan 03 in Wuhan, on Jan 06 in Hubei, and on Jan 23 in other provinces. (b) The impact of different timing of epidemic lockdown on the number of regional infections (upper row) and deaths (lower row) in Wuhan (left column), Hubei (middle column) and other provinces (right column) with social distancing activated on Jan 03 in Wuhan, on Jan 06 in Hubei, and on Jan 11 in other provinces. SARS-China infection: the infection number of the severe acute respiratory syndrome (SARS) in mainland China 2003, that is 5,779. MERS-global infection: the global infection number of Middle East Respiratory Syndrome (MERS) 2015, that is 2,269. SARS-China death: the death number of SARS in mainland China 2003, that is 829. MERS-global death: the global death number of MERS 2015, that is 431.

To put it more intuitively, we illustrated the estimated epidemic size and number of deaths in Wuhan, Hubei and other provinces with the optional strategies compared with the current strategy in Figure 4. Figure 4 (a) showed the estimated epidemic controlled with two of the theoretically best strategies in terms of minimizing the nationwide epidemic size and death (left column, epidemic lockdown on Jan 23 while social distancing activated in the whole nation simultaneously on Jan 03), and in terms of minimizing the epidemic outside the epicenter (right column, epidemic lockdown on Jan 03 while social distancing activated in the whole nation simultaneously on Jan 03). Figure 4 (b) showed the effects with two of the practically optional strategies aiming to minimize the nationwide epidemic size and death (left column, epidemic lockdown on Jan 23 while social distancing activated in Wuhan on Jan 03, in Hubei on Jan 06, in other provinces on Jan 11), and aiming to minimize the epidemic outside the epicenter (right column, epidemic lockdown on Jan 03 while social distancing activated in Wuhan on Jan 03, in Hubei on Jan 06, in other provinces on Jan 11). Figure 4 (c) showed the estimated epidemic controlled with the current strategy (simultaneously activated nationwide social distancing and epicenter lockdown on Jan 23) for comparison.

**Figure 4.**
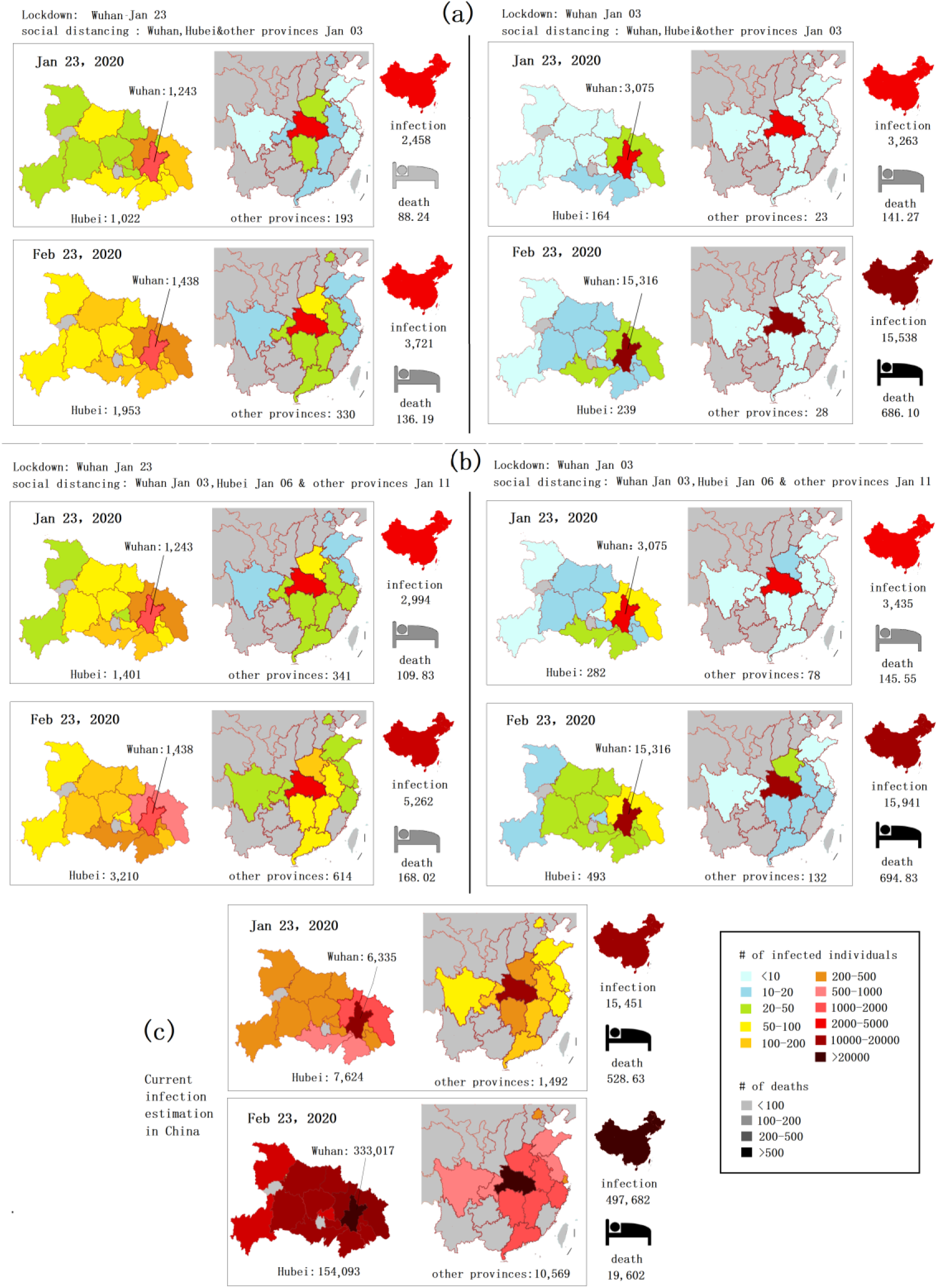
Illustrations of the estimated epidemic size and the number of deaths in Wuhan, Hubei and other provinces with the optional strategies compared with the current strategy. (a) Illustrations of the estimated epidemic with social distancing activated in the whole nation simultaneously on Jan 03. (b) Illustrations of the estimated epidemic with social distancing activated in Wuhan on Jan 03, in Hubei on Jan 06, in other provinces on Jan 11. The left column of (a) and (b): epicenter lockdown on Jan 23. The right column of (a) and (b): epicenter lockdown on Jan 03. (c) Illustrations of the estimated epidemic with current strategies: simultaneously activated nationwide social distancing and epicenter lockdown on Jan 23.

## Discussion

To improve the prevention and control of the COVID-19 epidemic, there are several questions concerning the effect of non-pharmaceutical interventions of social distancing and epicenter lockdown need to be addressed. We developed a data-driven SEIQR model of the COVID-19 epidemic in Wuhan, Hubei Province, and other provinces in China to investigate these questions.

The first question to be addressed is the timing to implement social distancing in the epicenter city, in other parts of the province, and the rest of the nation respectively. Our results showed that early intervention of social distancing could reduce the epidemic size significantly, especially synchronizing the intervention nation-widely at the possible earliest time. However, in such a large country like China, nationwide implementation of rigorous social distancing measures would bring profound socioeconomic influence. A stepwise intervention of social distancing as early as possible in the epicenter city, then in the province and later the whole nation would be more cost-effective.

The second question is the necessity to adopt social distancing to such a substantial extent. The intervention of social distancing in China was implemented in an all-of-Government and all-of-society manner. The specific measures were unprecedented comprehensive and rigorous, including but not limited to canceling gatherings, reducing public transportation, adopting enclosed community management, suspending work and school classes. Meanwhile, the socioeconomic cost was tremendous. Besides, these measures may be not universally practical in other countries. So we evaluated the impact of abolishing or reducing the extent of social distancing on the epidemic size. We found that moderate social distancing would remarkable attenuate its effect, while abolishing social distancing would leading to epidemic escalating.

The effectiveness of social distancing on epidemic control is still under academic debate. The results of virtual experiments vary. Some research suggested social distancing is effective, under the condition of early activation and long-lasting implement of combined measures [8], strict implement [6,9], and spontaneously adopted [10]; some concluded the effectiveness was mild [11]; some argued that moderate social distancing can worsen the disease outcome, [6,9] But real-world studies are rare. Here we quantified the impact of social distancing on the epidemic size in the real world, providing evidence that social distancing is effective to interrupt the transmission of the respiratory pathogen, especially with early implementation and to a substantial extent. Activating social distancing at the possible earliest time in the epicenter could gain time for the preparation of regions out of epicenter, and reduce wide range socioeconomic impact.

The third question is the necessity and the timing of the epicenter lockdown. Unexpectedly, our results showed that the significant decrease of the epidemic size by earlier social distancing would be partially neutralized by earlier epicenter lockdown. Further investigation suggested that the influences of epicenter lockdown on epidemic size and death differed between Wuhan and non-Wuhan regions. Earlier lockdown of Wuhan would deteriorate the situation in situ, but would largely lower the infection number in the rest of Hubei province and other provinces. The reasons may be as follows. After lockdown, a portion of the infected and the latently infected (the exposed in the model) would accumulate in Wuhan instead of exporting outside. Theoretically, it would not change the total infection or death number, but we found the conversion rate from the close contacts to the exposed is much higher in Wuhan than in other provinces, as listed in table S1 of Supplement Material, so is the death rate. Quickly surging infected patients in Wuhan exerted extraordinary pressure on local healthcare systems, resulting in an acute shortage of medical resources and healthcare workers. Then the fast-growing infection and resource shortage fueled each other in a vicious cycle. Numerous close contacts could not be quarantined for medical observation. Plentiful infected patients could not be hospitalized or quarantined for treatment. Then they would contact more susceptible persons and infect more people. Besides, unhospitalised and under-treated patients suffered a higher death rate. Taken together, the accumulation of patients in Wuhan caused by epicenter lockdown would aggravate the vicious circle. On the other hand, under the circumstances of nation-wide substantial social distancing, there were no such intense pressures on the healthcare systems out of Wuhan, so the conversion rate and death rate were much lower than those in Wuhan. Thus locking the epicenter down later, or even abolishing the lockdown, could reduce the infection density and the pressure of healthcare systems in the epicenter, and decrease the epidemic size and death to some extent. The price is that the epidemic size in the rest of the nation would be relatively larger than early lockdown, and the nationwide socioeconomic impact would be deepened. So it turns out to be a controversial issue to determine which option should be taken. To minimize the epidemic size and death, the epicenter city should not be locked down. To confine the epidemic distribution and mitigate the impact on nationwide socio-economy, the epicenter lockdown should be taken earlier.

There is little literature available concerning the impact of epicenter lockdown on the epidemic dynamics. Our results suggested a heterogeneous effect of epicenter lockdown on the epidemic size, death and localization. This founding could be partially evidenced by the epidemic prevalence on the Diamond Princess Cruise Ship. After COVID-19 appeared on the cruise ship, the whole ship was put on lockdown. A total of 3,711 passengers and crew were quarantined on the ship for 16-28 days. As of Mar 03, 2020, there were 706 infections and 6 death reported. The infection rate is up to 19%, and the death rate of 0.16%. The ship lockdown prevented epidemic spreading out, at the heavy price of within ship outbreak with an astonishingly high infection rate. The question then arises which aim should take the priority, minimizing the epidemic size and death, or better confining the prevalence to the origin region from spreading out. In addition, epidemics do more than cause death and debilitation. The regional socioeconomic damage after lockdown and the extensive socioeconomic impact of a nationwide spreading epidemic will further complicate the decision. This finding highlight the importance of careful evaluation and contemplation on the impact of epicenter lockdown from all side.

Taken together, the theoretically best strategy to minimize epidemic size and death is simultaneously activating nationwide substantial social distancing at the possible earliest time while abolishing epicenter lockdown. Alternatively, the ideal strategy to confine the epidemic within Hubei Province to the most extent is simultaneously activating both nationwide social distancing and epicenter lockdown at the possible earliest time. Practically, a stepwise implement of social distancing in Wuhan, Hubei and the rest of the nation would be more cost-effective.

Several issues to be noted when interpreting our work.

One is about the comprehensiveness of the interventions. In addition to the mainly discussed strategies of social distancing and epidemic lockdown, measures implemented in China also included forceful medical supports to Wuhan, reinforced quarantine management, and public health measures, e.g. requiring people to wear masks in public, raising public awareness for hand hygiene. Although these measures were not specifically discussed, their effectiveness has been illustrated by the adjustment of daily removed rate, and the decline of the base productive number *r0* in our model.

Another is about the time range of the interventions evaluated in this paper. We choose Jan 3, 2020 as the earliest available time point to implement interventions of social distancing and/or epicenter lockdown for two reasons. Practically, it is the date Wuhan started close contact management [1], a kind of preliminary quarantine management. Taking the move before this date could be better, but scarcely possible, so beyond the scope of our discussion. Technically, the available population migration data [7] covers the period from Jan 01 to Jan 22, so the timing of Wuhan lockdown investigated in this study is confined within this period. The effect of abolishing the epicenter lockdown was estimated under the assumption that the scale of the population migrated out of Wuhan would keep stable. Normally, the great migration departing big cities like Wuhan would pause around Jan 25, the day of Spring Festival, then the migration would continue with a reverse direction back to cities. However, under the circumstances of the epidemic prevailing, if the lockdown was abolished, individuals trying to get away from Wuhan might increase, leading to more imported cases in other regions than expected.

In addition, the application of our results should be tailored in a different situation. The non-Hubei provinces we studied were the ten provinces with most infections besides Hubei, plus the two municipalities of most importance in mainland China. These provincial regions had a large inflow of people from Wuhan, and had a high local population density. So there were more imported cases, and easy to trigger widespread local transmission. For those regions with little people flow from the epicenter and with a population of low density, it could be possible that reinforced quarantine management without stringent social distancing could be effective enough to interrupt local transmission. An extreme example is that there was only one COVID-19 case in Tibet Autonomous Region of China, and the one was an imported case from Wuhan. He was timely quarantined for treatment and discharged after recovery. 23 close contacts were quickly traced and quarantined for medical observation, and then ruled out of infection. For other regions in and out of China, the specific measures may be adjusted to the different social-political-economic environment.

## Conclusion

Based on the epidemic of COVID-19 in mainland China, we developed data-driven SEIQR models to investigate the impact of social distancing and epicenter lockdown on the epidemic dynamics. Activating social distancing at the possible earliest time with comprehensive and rigorous measures could significantly reduce the epidemic size of COVID-19. A stepwise implementation of social distancing in the epicenter city first, then in the province, and later the whole nation could be more practical and cost-effective. The decision of epicenter lockdown depends on the primary goal of epidemic control. To minimize the epidemic size and death, the epicenter city should not be locked down. To better confine the epidemic to the origin region, and mitigate the impact on nationwide socio-economy, the epicenter lockdown should be taken earlier.

Gratefully, the epidemic in mainland China has been declining steadily, especially in non-Wuhan regions. However, the epidemic out of China may just begin. Several countries, i.e. Korea, Italy, Iran, and Japan, etc. are confronted with an ongoing epidemic outbreak. With caution, the COVID-19 epidemic may even evolve into a global pandemic. Besides, other virulent infectious diseases may attack humans again in the future. We sincerely hope that our work could help the decision-making of epidemic prevention and control strategy for other countries in this COVID-19 epidemic and for future infectious disease epidemics. Tailored and sustainable approaches should be adopted in a different situation, striking a balance among the control of infection and death number, confining epidemic regions, and maintaining socioeconomic vitality.

## Data Availability

We are willing to share the data underlying the findings of our manuscript.

